# A sensitive and rapid wastewater test for SARS-COV-2 and its use for the early detection of a cluster of cases in a remote community

**DOI:** 10.1101/2021.08.13.21262039

**Authors:** Jade Daigle, Kathleen Racher, Justin Hazenberg, Allan Yeoman, Heather Hannah, Diep Duong, Umar Mohammed, Dave Spreitzer, Branden S. J. Gregorchuk, Breanne M. Head, Adrienne F.A. Meyers, Paul A. Sandstrom, Anil Nichani, James I. Brooks, Michael R. Mulvey, Chand S. Mangat, Michael G. Becker

**Author notes:** These authors contributed equally to this work (Chand S. Mangat, Michael G. Becker).

## Abstract

Throughout the COVID-19 pandemic, wastewater surveillance has been used to monitor trends in SARS-CoV-2 prevalence in the community. A major challenge in establishing wastewater surveillance programs, especially in remote areas, is the need for a well-equipped laboratory for sample analysis. Currently, no options exist for rapid, sensitive, mobile, and easy-to-use wastewater tests for SARS-CoV-2. The performance of the GeneXpert System, which offers cartridge-based, rapid molecular clinical testing for SARS-CoV-2 in a portable platform, was evaluated using wastewater as the input. The GeneXpert demonstrated a SARS-CoV-2 limit of detection in wastewater below 32 copies/mL with a sample processing time of less than an hour. Using wastewater samples collected from multiple sites across Canada during February and March 2021, a high overall agreement (97.8%) was observed between the GeneXpert assay and laboratory-developed tests regarding the presence or absence of SARS-CoV-2. Additionally, with the use of centrifugal filters the detection threshold of the GeneXpert system was improved to <10 copies/mL in wastewater. Finally, to support on-site wastewater surveillance, GeneXpert testing was implemented in Yellowknife, a remote community in Northern Canada where its use successfully alerted public health authorities to undetected transmission of COVID-19. The identification of SARS-CoV-2 in wastewater triggered clinical testing of recent travelers and identification of new COVID-19 cases/clusters. Taken together, these results suggest the GeneXpert is a viable option for surveillance of SARS-CoV-2 in wastewater in locations that do not have access to established testing laboratories.

## 1. Introduction

Since the start of the coronavirus disease 2019 (COVID-19) pandemic, public health officials worldwide have investigated methods to detect community transmission of the severe acute respiratory syndrome virus 2 (SARS-CoV-2). One such method includes environmental surveillance of wastewater, or wastewater-based epidemiology (WBE), that has been used for poliovirus monitoring (Asghar et al., 2014), illicit drug monitoring (Sulej-Suchomska et al., 2020), and detection of antimicrobial resistance (Hendriksen et al., 2019; Hutinel et al., 2019) amongst others. The observation that SARS-CoV-2 is shed in human stool (Chen et al., 2020; Holshue et al., 2020; Wölfel et al., 2020; Zhang et al., 2021) has led to renewed interest in WBE for COVID-19 surveillance and outbreak monitoring (Kitajima et al., 2020; Medema et al., 2020; Thompson et al., 2020; Tran et al., 2021). Detection of SARS-CoV-2 in wastewater is an early indicator of community transmission trends, often preceding clinical signals, such as new cases or hospitalizations, by 2-7 days (D’Aoust et al., 2021a; Peccia et al., 2020; Wurtzer et al., 2020). When combined with clinical data, SARS-CoV-2 prevalence in wastewater provides valuable insight to guide public health action. An additional advantage of WBE is that it is an equitable application of public health resources – it captures individuals who would be unlikely to present for clinical testing due to socioeconomic barriers or distrust of the medical community, and allows for community-wide surveillance in remote settings. Finally, WBE has also been used to monitor for potential outbreaks in high-risk institutional settings, such as long-term care facilities, correctional facilities, and dormitories (Accorsi et al., 2021; Davó et al., 2021; Harris-Lovett et al., 2021; Wang et al., 2020).

Despite these advantages, WBE testing typically requires a large laboratory due to the need for multiple types of instruments and equipment. Samples are often processed using lengthy and complicated protocols for concentration, extraction, and molecular testing that require specially trained individuals. Sample transport and shipping to centralized laboratories also causes delays, adding additional hours to days of sample processing time, negating its benefit as an early warning system (Larsen and Wigginton, 2020).

To date, no field-deployable and rapid test for SARS-CoV-2 in wastewater has been developed. Although a preprint article describes a rapid wastewater test from LuminUltra (Parra Guardado et al., 2020), this technology is not automated, requires basic laboratory skills, and currently has limited sensitivity data in real-world situations. The use of lateral flow immunological assays for SARS-CoV-2 in wastewater has been proposed (Hui et al., 2020; Orive et al., 2020; Tran et al., 2021) however, in clinical settings these assays display a low sensitivity when compared to the gold standard nucleic acid amplification tests (Jääskeläinen et al., 2021; Prince-Guerra et al., 2021). A possible solution for wastewater testing is the Cepheid GeneXpert system, which supports rapid, fully automated, cartridge-based clinical testing. Recently, Cepheid released a rapid diagnostic multiplex test with a run time of 37 minutes (Johnson et al., 2021), the Xpert^®^ Xpress-SARS-CoV-2/Flu/RSV combination test, for the detection of SARS-CoV-2, Influenza A, Influenza B, and Respiratory Syncytial Virus (RSV). This assay performs reverse transcription quantitative PCR (RT-qPCR) targeting the envelope (E) and nucleocapsid (N2) regions of the SARS-CoV-2 genome.

The GeneXpert has several characteristics that make it an ideal candidate for SARS-CoV-2 detection in wastewater as compared to other rapid diagnostic tests. The assay’s extraction step uses a filtration system that isolates and concentrates viral particles, while removing many of the inhibitors often present in wastewater. Moreover, the assay is one of the most sensitive rapid tests available, with a reported limit of detection of below 50 copies (cp)/mL in clinical settings (Becker et al., 2020; Johnson et al., 2021; Wolters et al., 2020; Zhen et al., 2020). The limit of detection of the GeneXpert can be further enhanced by monitoring the assay’s endpoint fluorescence, a practice used to improve sensitivity in clinical settings when performing high-multiplex sample pooling. Finally, the test is quantitative and provides a cycle threshold (CT) value that, through the use of a standard curve, can estimate the SARS-CoV-2 concentration in the sample.

To better track and anticipate COVID-19 disease trends, there is a need for an easy-to-use, mobile, and rapid wastewater test for SARS-CoV-2, particularly in remote communities or in resource-limited settings. Consequently, this study aimed to explore the use of the GeneXpert as a solution for SARS-CoV-2 testing in wastewater, which would allow for the decentralization of testing to sampling sites and the capacity to generate near real-time data to better guide public health actions.

## 2. Materials and Methods

### 2.1 Wastewater sample collection

Primary influent or raw wastewater samples were collected from two institutions, and 25 metropolitan and remote wastewater collections systems across Canada. Metropolitan sites included Toronto, Montreal, Vancouver, Edmonton, and Halifax, all samples from these sites were collected from mechanical wastewater treatment plants. Remote sites included five communities in the Northwest Territories: Yellowknife, Hay River, Inuvik, Fort Smith, and Fort Simpson. All samples from remote sites were collected from lift stations. Finally, two institutional samples were included in this work, each site houses between 300 to 800 long-term residents. All samples were composites collected in fresh polyethylene terephthalate bottles. All samples for this study were collected between February and May 2021. Samples that were transported for testing to PHAC-NML were shipped on ice packs and once received were refrigerated at 4°C for up to 48 hours until testing was performed.

### 2.2 Preparation of wastewater samples for GeneXpert testing

Two methods were used to prepare wastewater samples for processing on the GeneXpert System. These methods are referred to as <u>Method A</u> (no concentration) and <u>Method B</u> (concentration of supernatant via Amicon centrifugal filtration).

#### Method A

15 µL of 10% Tween 80 [final 0.01% (v/v)] was added to 15 mL of untreated wastewater. The sample was then vortexed at maximum speed for 20 seconds and then rested for two minutes to allow debris to settle. Once settled, 300 µL of sample was loaded directly into the GeneXpert cartridge and analyzed as per manufacturer’s protocol for clinical specimens.

#### Method B

15 µL of 10% Tween 80 [final 0.01% (v/v)] was added to 15 mL of wastewater and vortexed at maximum speed for 20 seconds. The wastewater was then clarified via centrifugation for 20 minutes at 4,200 x g and 4°C. The supernatant was decanted into an Amicon^®^ Ultra-15 10 kDa Centrifugal Filter Unit (MilliporeSigma, Burlington, MA), and care was taken to avoid dislodging of the pellet. The Amicon filtration devices were centrifuged at 4,200 x g for 30 minutes and 4°C. Molecular grade water was added to concentrates to bring the volume to 300 µL (input volume for GeneXpert testing). Next, the sample was loaded directly into the GeneXpert cartridge and analyzed as per the manufacturer’s protocol for clinical specimens.

### 2.3 Mobile wastewater testing for SARS-CoV-2 with the GeneXpert System

All wastewater samples were tested on the GeneXpert^®^ XVI (Cepheid, Sunnyvale, CA) using the Xpert^®^ Xpress-SARS-CoV-2/Flu/RSV cartridge (Cepheid, Sunnyvale, CA). GeneXpert tests were performed at PHAC-NML in Winnipeg, Canada and the Taiga Environmental Laboratory in Yellowknife, Canada. The endpoint fluorescence was also monitored for each run, where an endpoint fluorescence of >10 is indicative of a weakly positive sample (reported to public health as a trace detection). The use of GeneXpert endpoint fluorescence to capture weak positives is a practice used in some Canadian hospitals to determine if pooled samples should be split for individual testing. For concentrated wastewater samples (<u>Method B</u>), SARS-CoV-2 concentration was determined using a standard curve (Becker et al., 2020; Johnson et al., 2021b). To conserve GeneXpert supplies for clinical testing, all initial experiments were performed with unused cartridges from open test kits that were sampled as part of lot quality management.

### 2.4 Preparation of wastewater nucleic acid extracts for conventional laboratory testing

PHAC-NML has protocols to process both the supernatant and solid fractions of wastewater (Peterson et al., 2021). Samples received from Northwest Territories and Vancouver underwent both the supernatant and solids processing methods, while only the solids were processed for samples received from Toronto, Montreal, Edmonton, and Halifax wastewater treatment plants as per the preferences of these communities.

The method used to process the supernatant followed the same concentration steps outlined in <u>Method B</u>, with the exception of a filtration centrifugation time of 35 minutes. After filtration, 2 mg of carrier RNA (Sigma-Aldrich, St. Louis, MO) and 700 µL of MagNA Pure 96 External Lysis Buffer (Roche, Pleasanton, CA) was added to the processed supernatant before extraction on the MagNA Pure 96 Instrument (Roche, Pleasanton, CA).

For processing the solids fraction, 30 mL of wastewater was centrifuged for 20 minutes at 4200 x g and 4°C. After centrifugation, the supernatant was carefully removed and the pellet was transferred to a bead beating tube containing 200 µL of 0.5 mm zirconia/silica beads and 700 µL of Buffer RLT lysis buffer with 1% β-mercaptoethanol (Qiagen, Germantown, MD). Samples were homogenized using a Bead Mill 24 Homogenizer (Fisher Scientific, Hampton, NH) for 4 cycles of 30 seconds at 6m/s with 20 seconds of rest time between each cycle. Subsequently, the homogenate was clarified at 12000 x g for 3 min, and 1000 µL of the clarified homogenate was transferred to a 96-well MagNA Pure 96 Processing Cartridge containing 2 mg of carrier RNA. The extraction was then performed on the MagNA Pure 96 Instrument (Roche, Pleasanton, CA).

### 2.5 Laboratory-based wastewater testing for SARS-CoV-2 by RT-qPCR

SARS-CoV-2 was quantified in 5 µL of lysate using the TaqPath™ 1-Step RT-qPCR Master Mix, CG (Life Technologies, Carlsbad, CA) with N1 and N2 primer and probe sequences obtained from the US Centers for Disease Control and Prevention (Supplementary Table 1). The RT-qPCR reaction was prepared as per the manufacturer’s instructions with a primer concentration of 500 nM and probe concentration of 125 nM. The following cycling conditions were used: 25°C for 2 minutes; 50°C for 15 minutes; 95°C for 2 minutes; followed by 40 cycles of 95°C for 5 seconds and 60°C for 30 seconds. RT-qPCR was performed with two technical replicates. Quantification of the SARS-CoV-2 N1 and N2 CT values was performed using a 5-point standard curve prepared from synthetic DNA oligos. All TaqMan-based RT-qPCR assays were performed on a QuantStudio™ 5 Real-Time PCR System (Life Technologies, Carlsbad, CA).

### 2.6 Standardization of SARS-CoV-2 Measurements

High-titre SARS-CoV-2 culture (Strain VIDO; GISAID Accession: EPI_ISL_425177), made inactive by gamma-irradiation, was provided by the Special Pathogens Program of PHAC-NML. Briefly, SARS-CoV-2 was cultured in Vero cells in minimum essential media, and cellular debris was removed via low-speed centrifugation. Viral culture was quantified using the GeneXpert system and standard curve, as well as through droplet digital PCR.

## 3. Results

### 3.1 Detection of gamma-irradiated SARS-CoV-2 culture in wastewater

To determine if the GeneXpert SARS-CoV-2 test was compatible with wastewater, gamma-irradiated SARS-CoV-2 was serially diluted in wastewater that was negative for SARS-CoV-2 as determined by the laboratory-developed wastewater test of the Public Health Agency of Canada National Microbiology Laboratory Winnipeg (See Materials and Methods). At input concentrations of 32, 64, and 128 cp/mL the assay detected SARS-CoV-2 in all replicates (supplemental Figure 1). All replicates were negative at input concentrations of 16 cp/mL.

### 3.2 SARS-CoV-2 detection is robust in unconcentrated wastewater samples from Canadian wastewater treatment plants

The GeneXpert was used to test wastewater collected in February and March of 2021 from various Canadian communities (Method A, Materials and Methods). In total, 30 samples were collected from communities with active COVID-19 cases, and 15 samples from were collected from communities with no known COVID-19 activity (Figure 1). The identity of the specimens was censored until test results were collated. One sample produced a loading error on the GeneXpert system, possibly due to clogging of the test cartridge; however, a repeat test was performed successfully. All samples were tested concurrently with our laboratory-developed test for COVID-19 targeting the solids fraction of wastewater, and for a subset, the supernatant fraction was also tested. Our laboratory-developed tests detected SARS-CoV-2 in all 30 wastewater samples taken from communities with active cases of COVID-19 (Figure 1). The viral concentration ranging from 9.2 to 216 cp/mL (solids fraction). Of these positives, 22 were reported as positive on the GeneXpert, seven were endpoint positive (weak positive), and one was negative for an agreement of 96.6%. As expected, our laboratory-developed test did not detect SARS-CoV-2 in any of the wastewater samples collected from communities with no active cases of COVID-19 (n = 15). The GeneXpert assay also reported all of these samples as negative for SARS-CoV-2 (negative agreement of 100%).

**Figure 1:**
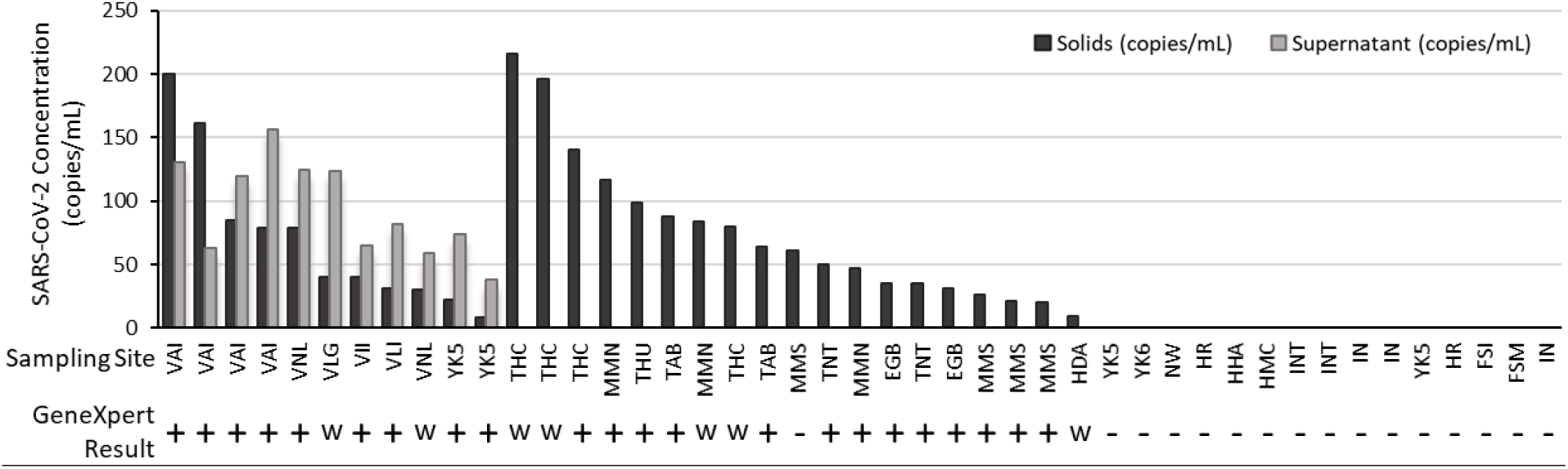
SARS-CoV-2 concentration in Canadian wastewater samples, and GeneXpert^®^ rapid test results. SARS-CoV-2 concentration was determined using a laboratory-developed solids-based extraction and RT-qPCR test targeting N1 and N2. Value shown is the average of N1 and N2 targets. For a subset of samples, SARS-CoV-2 concentration was measured in the liquid fraction with a laboratory-developed test. Locations listed multiple times refer to unique samples collected on different days. All 15 samples on the far right of the graph (negative for SARS-CoV-2) were collected from communities with no active SARS-CoV-2 cases. GeneXpert results are either positive (+), weakly positive as determined by endpoint fluorescence (w), or negative (-).Legend: EGB = Edmonton Goldbar; HAD = Halifax Dartmouth; HHA = Halifax Halifax; HMC = Halifax Millcove; MMN = Montreal North; MMS = Montreal South; TAB = Toronto Ashbridges Bay; THC = Toronto Highland Creek; THU = Toronto Humber; TNT = Toronto North Toronto; VAI = Vancouver Annacis Island; VII = Vancouver Iona Island; VLG = Vancouver Lions Gate; VLI = Vancouver Lulu Island; VNL = Vancouver Northwest Langley; YK5 = Yellowknife Lift Station 5; YK6 = Yellowknife Lift Station 6; HR = Hay River; FSM = Fort Smith; FSI = Fort Simpson; IN = Inuvik; INT = Institutional sample; NW = Norman Wells Sewer.

### 3.3 Wastewater concentration increases GeneXpert test sensitivity in communities with a low prevalence of SARS-CoV-2

Concentration of samples via Amicon centrifugal filters (Method B, materials and methods) was used to improve the sensitivity of the GeneXpert rapid test and to investigate if this would allow the test to be used quantitatively (without concentration, the assay’s input of 300 µL makes quantification unreliable due to sampling biases and the heterogeneous nature of wastewater). Eleven wastewater samples were selected from the PHAC Wastewater Surveillance Program with concentrations between 8.5 to 273 cp/mL, and tested on the GeneXpert system with or without concentration via centrifugal filtration. Two wastewater samples (YK5-2, YK5-3) were only positive for SARS-CoV-2 when samples were concentrated with centrifugal filters (Figure 2A). Overall, CT value decreased by an average of 3.9 cycles, or approximately 16-fold, with the use of centrifugal filters. Following concentration via centrifugal filtration, one sample from a major urban centre also reported as positive for Influenza A (data not shown).

**Figure 2:**
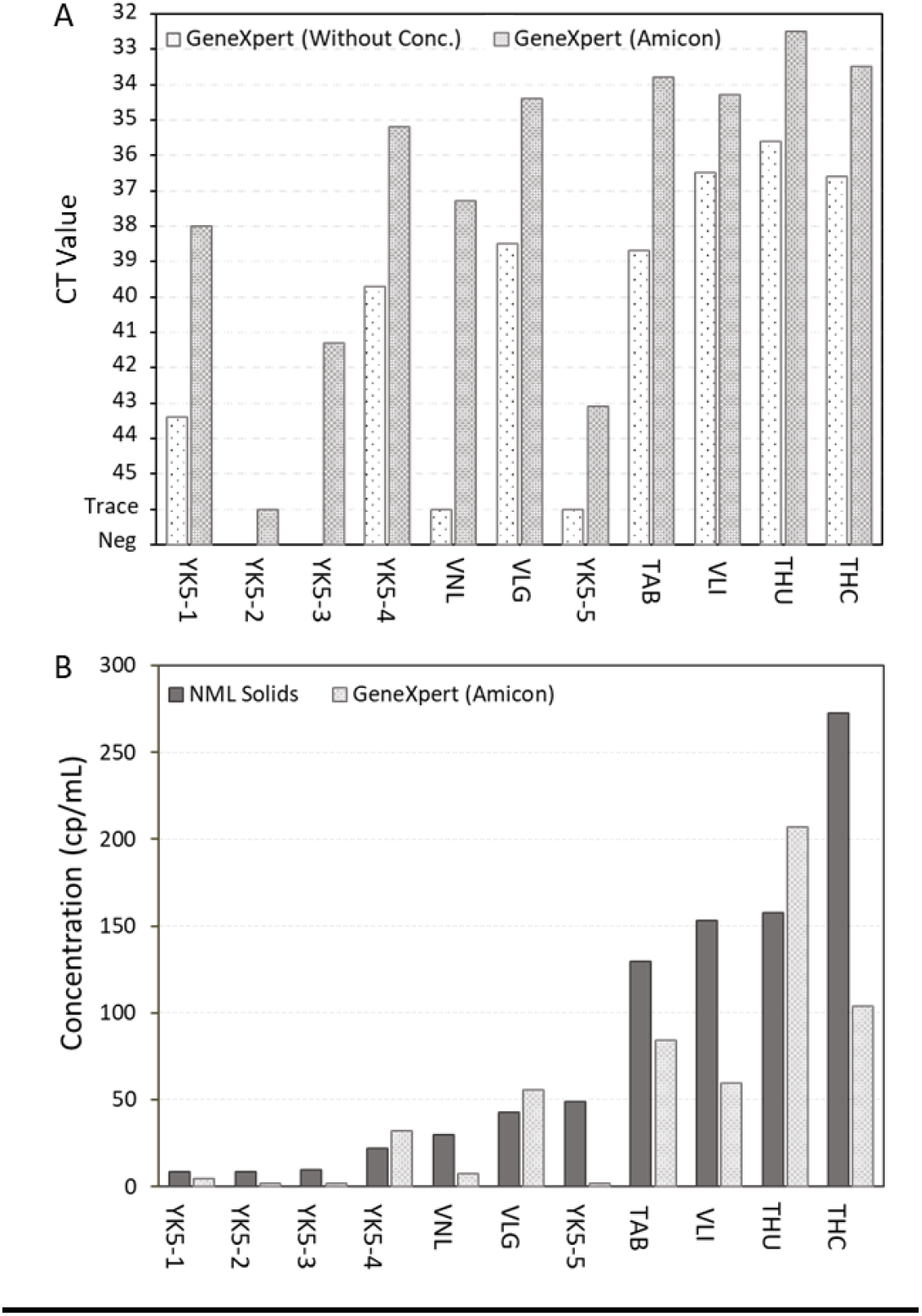
Use of Amicon centrifugal filters to increase sensitivity of the GeneXpert^®^ SARS-CoV-2/Flu/RSV assay. (A) The effect of centrifugal filtration on GeneXpert CT values. Left white bars show CT values without the use of centrifugal filters, right grey bars show values with the use of centrifugal filters. (B) Quantification of SARS-CoV-2 concentration in wastewater using centrifugal filters and GeneXpert standard curve (light grey), as compared to results from laboratory-developed solids assay (dark grey). Legend: TAB = Toronto Ashbridges Bay; THC = Toronto Highland Creek; THU = Toronto Humber; VLG = Vancouver Lions Gate; VLI = Vancouver Lulu Island; VNL = Vancouver Northwest Langley; YK5 = Yellowknife Lift Station 5 (multiple time points).

A standard curve was developed to convert CT values into viral load in previous work using the GeneXpert for clinical testing (Becker et al., 2020; Johnson et al., 2021). This same curve was used here to convert GeneXpert CT values into SARS-CoV-2 concentration in wastewater. GeneXpert SARS-CoV-2 concentration was then compared to estimates from the PHAC-NML laboratory-developed solids assay (Fig. 2B). There was a moderate/strong correlation (r = 0.724) between the predicted SARS-CoV-2 concentrations of the GeneXpert and the laboratory-developed solids assay (Fig. 2B).

### 3.4 The GeneXpert SARS-CoV-2 assay can serve as an early warning system for COVID-19 in remote communities without known cases

The PHAC National Wastewater Surveillance Program has been testing wastewater in the Northwest Territories since August 2020. Wastewater samples were shipped from the Northwest Territories to PHAC-NML in Winnipeg for testing with a transit time of 1-4 days, with a further two days required for testing and reporting. In March 2021, a GeneXpert system was deployed to Yellowknife to support an on-site wastewater surveillance program and expedite test reporting. GeneXpert surveillance in Yellowknife formally began on March 26 with wastewater tested multiple times weekly (Fig 3).

**Figure 3:**
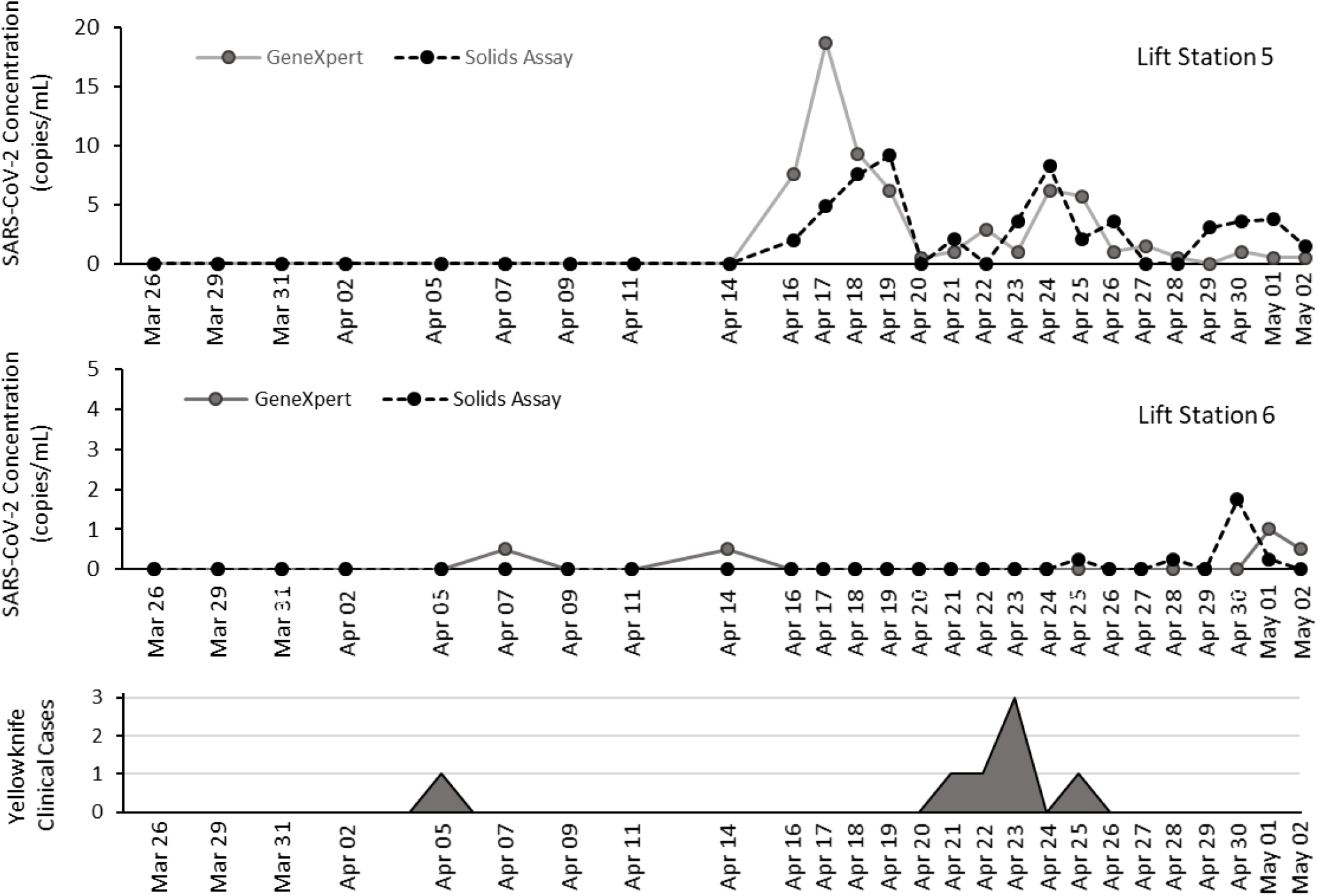
Wastewater surveillance of two Yellowknife lift stations with the GeneXpert and laboratory-developed solids assay. (A) Measured SARS-CoV-2 concentration in Lift Station 5 wastewater using the GeneXpert-Amicon rapid test (solid grey line), or laboratory-developed solids assay at the National Microbiology Laboratory (dotted black line). Collection frequency increased after first detection. (B) Measured SARS-CoV-2 concentration in wastewater collected at Lift Station 6. (C) New cases identified in Yellowknife between March 26 – May 2. One travel-related case was identified on April 6, and a second independent cluster was identified on April 21-25.

Preceding the GeneXpert pilot study initiated on March 26, wastewater samples from Yellowknife were shipped to and tested exclusively at the PHAC-NML laboratory in Winnipeg, Canada, a distance of approximately 1745 kilometres (Fig 4A). These wastewater samples were collected from two major lift stations that capture wastewater from >85% of the Yellowknife population (Fig 4B).

**Figure 4:**
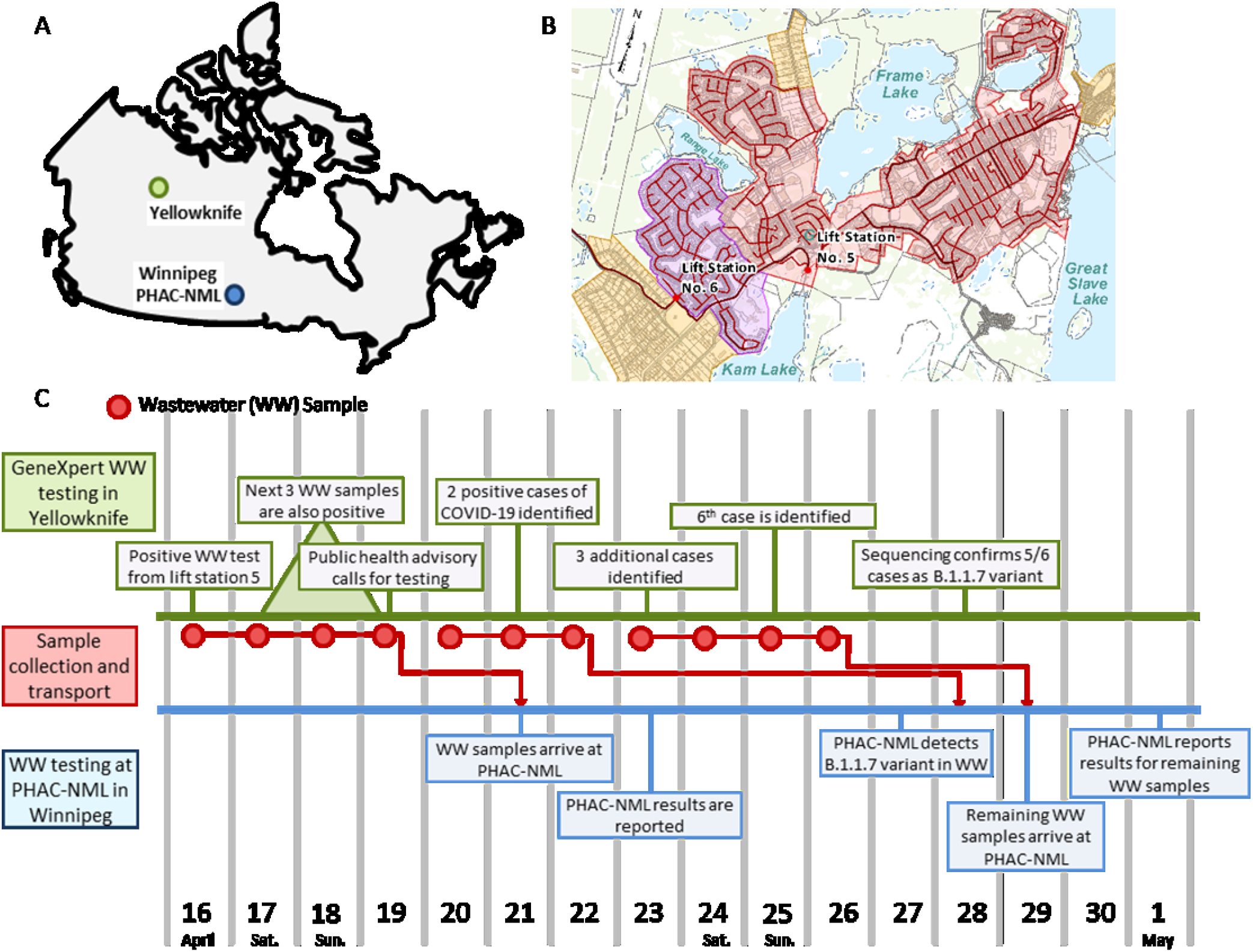
Timeline of events during the GeneXpert^®^ wastewater surveillance pilot in Yellowknife, Canada. (A) Map showing locations of Yellowknife (green) and PHAC-NML laboratory in Winnipeg (blue). (B) Two main lift stations (No. 5 and No. 6) in Yellowknife, and their corresponding catchments covering >85% of the Yellowknife population. (C) Timeline of events leading to the identification of a SARS-CoV-2 outbreak in Yellowknife. Timing of wastewater test results and public health actions in Yellowknife are shown in green, sample collection and transport events are shown in red, and at PHAC-NML testing is shown in blue.

Two weak positives (based on endpoint fluorescence) were observed on the GeneXpert system on April 7 and April 14 (Figure 3B) in Lift Station 6 coinciding with a travel-related case that was reported on April 5 (Figure 3C). Verbal communication with Northwest Territories’ Office of the Chief Public Health Officer acknowledged the wastewater signal from Lift Station 6 was consistent with information from clinical investigations.

Beginning with samples collected from Lift Station 5 on April 16, both the GeneXpert, and subsequently also the PHAC-NML assay, detected multiple SARS-CoV-2 signals in Yellowknife wastewater. As a response to the first detection of SARS-CoV-2, wastewater collection in Yellowknife was adjusted to daily sampling. Three weeks after initiating the GeneXpert pilot, a persistent SARS-CoV-2 signal was detected in wastewater. Four consecutive positive wastewater detections triggered the Office of the Chief Public Health Officer to initiate a response, including recommending COVID-19 testing for recent travellers to the NWT. This testing and subsequent contact tracing led to the identification of a cluster of six cases in the community during the period of April 20-26 (Fig 3C). The timeline for this response is summarized in Figure 4C.

### 3.5 Yellowknife GeneXpert wastewater surveillance pilot

Yellowknife wastewater samples were also shipped to PHAC-NML for confirmatory testing. The first batch of wastewater samples were received by PHAC-NML on April 21; however, results were reported on April 23, several days after the first Yellowknife COVID-19 cases had been already identified. Results from the PHAC-NML assay were consistent with the data produced by the GeneXpert. As part of the laboratory-developed assay, information is also provided on the variants present in the wastewater sample (Peterson et al., 2021). PHAC-NML reported B.1.1.7 (alpha variant) as the majority variant present in Yellowknife wastewater. This was consistent with clinical sequencing data, which identified most cases in the cluster as B.1.1.7.

## 4. Discussion

Currently, no options exist for mobile and rapid testing of wastewater. This study explored the GeneXpert system as a candidate technology for this purpose, and performed a pilot study in Yellowknife, Canada. The data presented here demonstrates that the GeneXpert SARS-CoV-2/Flu/RSV assay can reliably detect SARS-CoV-2 in wastewater at concentrations above 32 cp/mL (Figure S1; Figure 1). Sensitivity of the assay can be improved further through concentration methods such as centrifugal filtration, which facilitated detection of SARS-CocV-2 below 10 cp/mL (Fig 2B). Although effective, concentration by centrifugal filters is not ideal, as it requires use of a centrifuge, expensive specialized filters, and additional processing time by the operator. Future work should investigate more rapid and deployable methods for concentration, such as filter syringes or concentrating pipettes (Gonzalez et al., 2020). Alternatively, concentration of the sample may not be required if the GeneXpert system is used to monitor wastewater from a smaller system (small neighborhood or institutional samples).

At the observed level of sensitivity, the GeneXpert is capable of serving as an early detection system in remote communities. This is supported by data from the Yellowknife pilot project, where the GeneXpert system detected SARS-CoV-2 in wastewater before public health officials were aware of its presence in the community. Previously, Yellowknife relied exclusively on wastewater testing at a Winnipeg-based testing facility (PHAC-NML), which delayed results by approximately 4-7 days due to shipping and sample processing time. In the time between sample collection in Yellowknife and completion of testing in Winnipeg, seven positives wastewater samples were recorded on the GeneXpert system and five COVID-19 cases were already identified in the community. Since this pilot, the Government of Northwest Territories has further expanded GeneXpert testing to include seven additional remote communities. Together, this clearly demonstrates the utility of rapid, deployable wastewater testing for SARS-CoV-2.

In terms of efficient resource allocation, a single GeneXpert cartridge can effectively screen an entire community with much broader coverage than a single clinical test, and is likely a more sustainable and cost-effective option for community surveillance in post-vaccination scenarios. Additionally, because of the widespread distribution GeneXpert systems for SARS-CoV-2, HIV, tuberculosis, and influenza testing, this technology provides a means to improve global access to wastewater surveillance. This includes remote Canadian communities as well as remote areas of South America, Africa, and Asia.

There are some limitations to the GeneXpert technology. Notably, the targets are determined by the manufacturer and the system itself is optimized for clinical testing. The system currently does not monitor for SARS-CoV-2 variant detection, nor include fecal indicators such as pepper mild mottle virus that may be useful for data normalization (D’Aoust et al., 2021b). As the cartridge uses an internal filter there is also a risk of clogging; however, this was only observed once in this study and the sample was successfully processed in a repeat test. The input volume of the GeneXpert SARS-CoV-2/Flu/RSV test is also quite low at 300 µL, which may introduce sampling biases when testing large volumes of heterogeneous wastewater. This sampling bias makes any quantitative data from the GeneXpert less reliable unless a concentration step is performed. Ideally, wastewater-specific test cartridges for the GeneXpert could be developed with optimized targets and larger input volumes; or a similar large-volume assay could be developed using a competing technology.

The Xpert Xpress SARS-CoV-2/Flu/RSV assay used in this study is also used clinically for the detection of Influenza A, Influenza B, and Respiratory Syncytial Virus. Therefore, it is theoretically possible that this assay will be able to detect these other pathogens in wastewater in addition to SARS-CoV-2. In this study, we only observed one positive detection of Influenza A in a wastewater sample from a major Canadian city, and no detection of Influenza B or Respiratory Syncytial Virus. The lack of detection may be reflective of the low transmission rates of influenza and other respiratory viruses in 2020-2021 (Huang et al., 2021; Lagacé-Wiens et al., 2021; Lee et al., 2021), or may indicate the GeneXpert is not suitable for surveillance of these pathogens warranting further studies. Other clinical tests for the GeneXpert system have targets likely to be present in wastewater such as norovirus, *Clostridium difficile*, and various antimicrobial resistance genes. Of particular important in remote and isolated communities would be surveillance for active Tuberculosis. It would also be of interest to investigate if these clinical GeneXpert assays are also compatible with wastewater as an input.

Taken together, this work highlights the importance of rapid wastewater testing for SARS-CoV-2. Although previous studies have shown wastewater signals may provide several days of lead time as compared to clinical cues of transmission, much of this early warning is eroded by delays resulting from sample shipping and processing times. Centralized laboratories in urban centres often provide wastewater surveillance for remote and isolated communities; however, these locations are often separated by large distances creating logistical complications. As the second largest country by land mass, wastewater tests will need to be deployable for an effective and equitable Canadian national wastewater surveillance program for COVID-19. Additionally, WBE has not been substantially applied to low-resource communities (Naughton et al., 2021). Vaccinations and effective clinical surveillance has not reached many parts of the developing world, and these factors increase the risk and impact from the introduction of COVID-19 into these communities (Ritchie et al., 2020).

## 5. Conclusions

In conclusion, the GeneXpert can be used for wastewater testing for SARS-CoV-2, improving access to wastewater based epidemiology, and providing rapid results allowing for an immediate public health response. In particular, this technology could be applied in support wastewater surveillance in remote communities where it may provide early warning of SARS-CoV-2 outbreaks. It is likely wastewater surveillance will become increasingly important in the post-vaccination pandemic landscape as individuals with asymptomatic/mild infections continue transmitting SARS-CoV-2 but are unlikely to be clinically tested. Although not specifically designed for this purpose, the GeneXpert system provides a stopgap measure until a purposely-designed wastewater rapid test for SARS-CoV-2 becomes available for remote and low-resource communities. Ideally, future solutions should provide real-time or near real-time monitoring of SARS-CoV-2 and other pathogens in wastewater to inform public health responses.

## Data Availability

All data are available in the manuscript.

## 6. Acknowledgements

We would like to thank all members of the PHAC National Wastewater Surveillance Program for their technical assistance and support. We also thank the Centre for Population Health Data of Statistics Canada (Audra Nagasawa) for coordination of wastewater sampling to support the National Wastewater Surveillance Program. We also want to acknowledge the Office of the Chief Public Health Officer (Dr. Andy Delli Pizzi and Dr. Kami Kandola) for their work translating positive wastewater signals into appropriate public health action.

## 9. Supplementary Material

**Supplementary Table 1:**
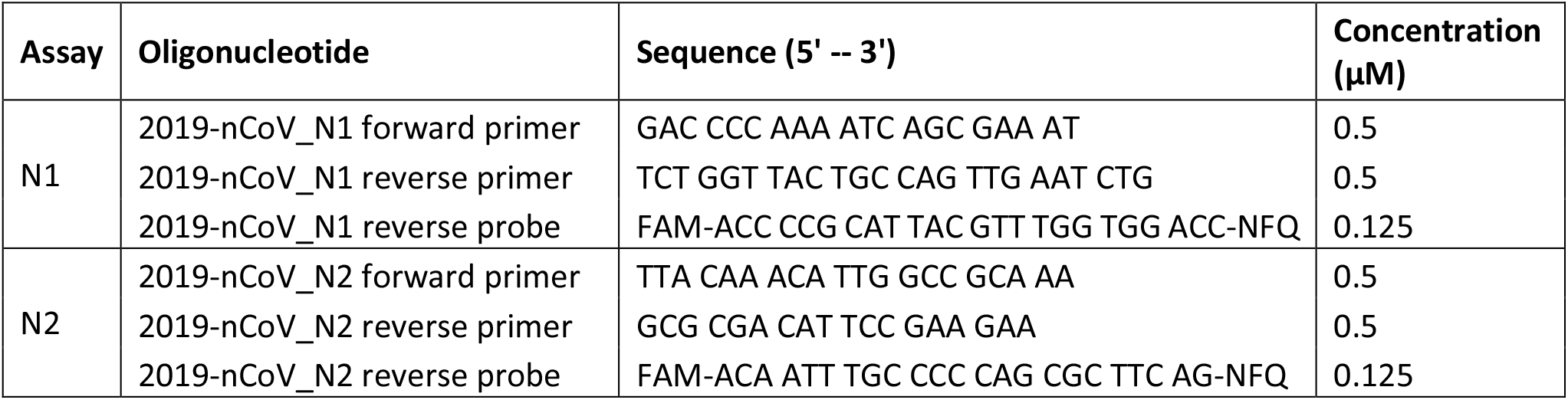
List of SARS-CoV-2 Primers and Probes used for the Laboratory-Developed q-RT-PCR Test.

**Supplementary Figure 1:**
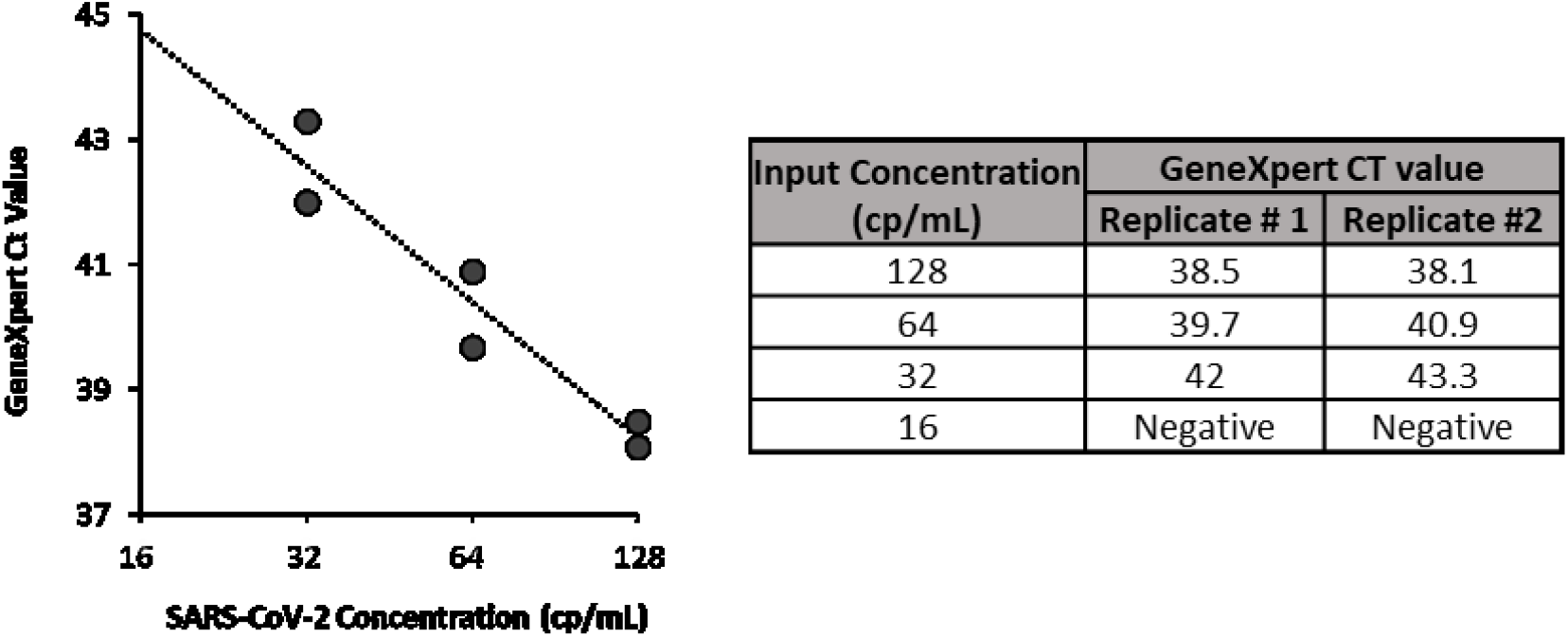
Detection of SARS-CoV-2 in wastewater using the GeneXpert system. Gamma irradiated SARS-CoV-2 culture fluid was diluted in wastewater to final concentrations between 16 – 128 copies (cp/mL). Tests were performed in duplicate. The GeneXpert detected SARS-CoV-2 in all replicates with an input equal to or greater than 32 cp/mL.

## Notes

### Competing Interest Statement

The authors have declared no competing interest.

### Clinical Trial

This study did not involve clinical specimens.

### Funding Statement

Experiments were funded and supported by the Public Health Agency of Canada and Government of the Northwest Territories.

### Author Declarations

This study did not involve clinical specimens, and tested environmental/wastewater samples for SARS-CoV-2.

